# A single-cell atlas of the peripheral immune response to severe COVID-19

**DOI:** 10.1101/2020.04.17.20069930

**Authors:** Aaron J. Wilk, Arjun Rustagi, Nancy Q. Zhao, Jonasel Roque, Giovanny J. Martinez-Colon, Julia L. McKechnie, Geoffrey T. Ivison, Thanmayi Ranganath, Rosemary Vergara, Taylor Hollis, Laura J. Simpson, Philip Grant, Aruna Subramanian, Angela J. Rogers, Catherine A. Blish

## Abstract

There is an urgent need to better understand the pathophysiology of Coronavirus disease 2019 (COVID-19), the global pandemic caused by SARS-CoV-2. Here, we apply single-cell RNA sequencing (scRNA-seq) to peripheral blood mononuclear cells (PBMCs) of 7 patients hospitalized with confirmed COVID-19 and 6 healthy controls. We identify substantial reconfiguration of peripheral immune cell phenotype in COVID-19, including a heterogeneous interferon-stimulated gene (ISG) signature, HLA class II downregulation, and a novel B cell-derived granulocyte population appearing in patients with acute respiratory failure requiring mechanical ventilation. Importantly, peripheral monocytes and lymphocytes do not express substantial amounts of pro-inflammatory cytokines, suggesting that circulating leukocytes do not significantly contribute to the potential COVID-19 cytokine storm. Collectively, we provide the most thorough cell atlas to date of the peripheral immune response to severe COVID-19.

## INTRODUCTION

Coronavirus disease 2019 (COVID-19), caused by severe acute respiratory syndrome coronavirus 2 (SARS-CoV-2), has become a global pandemic, with more than 2 million cases worldwide (as of April 15, 2020) since the first reports of the disease in December 2019. Approximately 20% of patients with COVID-19 develop severe disease warranting hospitalization and approximately 5% require intensive care^1^. The incidence of severe COVID-19 increases with age, as well as underlying conditions including emphysema, obesity, diabetes, and hypertension^2,3^.

Severe COVID-19 is associated with increased levels of interleukin (IL)-6, IL-10 and tumor necrosis factor (TNF)-α^4,5^. Whether higher levels in more severely ill patients reflect an appropriate response to more severe disease or whether these reflect a dysregulated immune response, often described as a “cytokine storm,” is unknown. Recent studies have demonstrated a population of peripheral inflammatory monocytes producing high levels of cytokines in patients with severe COVID-19^6,7^. In addition, lymphopenia is frequently observed in severe COVID-19, particularly a reduction in CD4^+^ and CD8^+^ T cells, and remaining T cells have a less functional and more exhausted phenotype^8,9^. Inflammatory immune cell infiltrates may contribute to lung pathology in COVID-19^10^, similar to SARS^11,12^. To elucidate pathways that might lead to immunopathology or protective immunity in severe COVID-19 across all immune cell subsets in peripheral blood, we used single-cell RNA sequencing (scRNA-seq) to profile peripheral blood mononuclear cells (PBMCs) from 7 patients hospitalized for COVID-19 and 6 healthy controls.

## RESULTS

### Single-cell transcriptomic profiling captures shifts in peripheral immune cell composition in SARS-CoV-2 infection

To profile the peripheral immune response to severe COVID-19, we performed Seq-Well-based^13^ massively parallel scRNA-seq on 8 peripheral blood samples from 7 hospitalized patients with RT-PCR-confirmed SARS-CoV-2 infection and 6 healthy controls. The demographic characteristics, clinical features, and outcomes of these patients are listed in **Table 1**. The seven subjects profiled were all male and ranged in age from 20 to >80 years of age and samples were collected between two and sixteen days following onset of symptoms. Four of eight COVID-19 samples were collected from patients who were ventilated and diagnosed with acute respiratory distress syndrome (ARDS); the remaining four samples were from less severely ill patients (**Table 1**). One patient (C1) was sampled twice, initially at 9 days post-symptom onset while hospitalized and requiring supplemental oxygen but not ventilated, and again at 11 days post-symptom onset following intubation. At the time of analysis, only subject C6, the eldest patient, was deceased; subjects C2, C6, and C7 had been discharged home, and the remainder were still in the hospital (**Table 1**). Five patients received remdesivir as part of a separate trial. Demographic characteristics of healthy donors are reported in **Extended Data Table 1**.

**Table 1:** Demographics, sample characteristics, and disease course of COVID-19 patients.

| Donor | Age | Sex | Days from reported symptom onset | Days from closest reported or measured fever | Admission level | Ventilated/ARDS? | Days intubated | Days paralyzed | PaO<sub>2</sub>/FI0<sub>2</sub> on ICU admission | Clinical outcome |
| --- | --- | --- | --- | --- | --- | --- | --- | --- | --- | --- |
| C1 | 60-69 | M | 9 | 9 | ICU | No |  |  |  | Now extubated, on room air, expect discharge home |
|  |  |  | 11 | 11 |  | Yes | 17 | 2 | 91 |  |
| C2 | 40-49 | M | 16 | 16 | ICU | No | - | - | - | Discharged home |
| C3 | 30-39 | M | 9 | 9 | ICU | Yes | 25 | 4 | 100 | Tracheostomy, prolonged ICU course |
| C4 | 30-39 | M | 9 | 9 | ICU | Yes | 16 | 3 | 71 | Now extubated, on supplemental oxygen, expect discharge home |
| C5 | 50-59 | M | 15 | 1 | ICU | No | - | - | - | Discharged home |
| C6 | >80 | M | 2 | No fever | ICU | Yes | 11 | 0 | 126 | Deceased |
| C7 | 20-29 | M | 12 | No fever | Floor | No | - | - | - | Discharged home |

In total, we sequenced 44,271 cells with an average of 3,194 cells per sample. We created a cells-by-genes expression matrix of all cells which we used to perform dimensionality reduction by Uniform Manifold Approximation and Projection (UMAP) and graph-based clustering, which identified 30 clusters of cells (**Fig. 1a**). Next, we calculated each cluster’s most highly differentially expressed (DE) genes to manually annotate clusters with their respective cellular identities (**Fig. 1b, c; Methods)**, and confirmed these identities using SingleR^14^ for automated reference-based annotation (**Extended Data Fig. 1**). The dimensionality reduction indicated substantial phenotypic differences between COVID-19 cases and controls, predominantly in monocytes, T cells, and natural killer (NK) cells (**Fig. 1a, b**).

**Figure 1.**
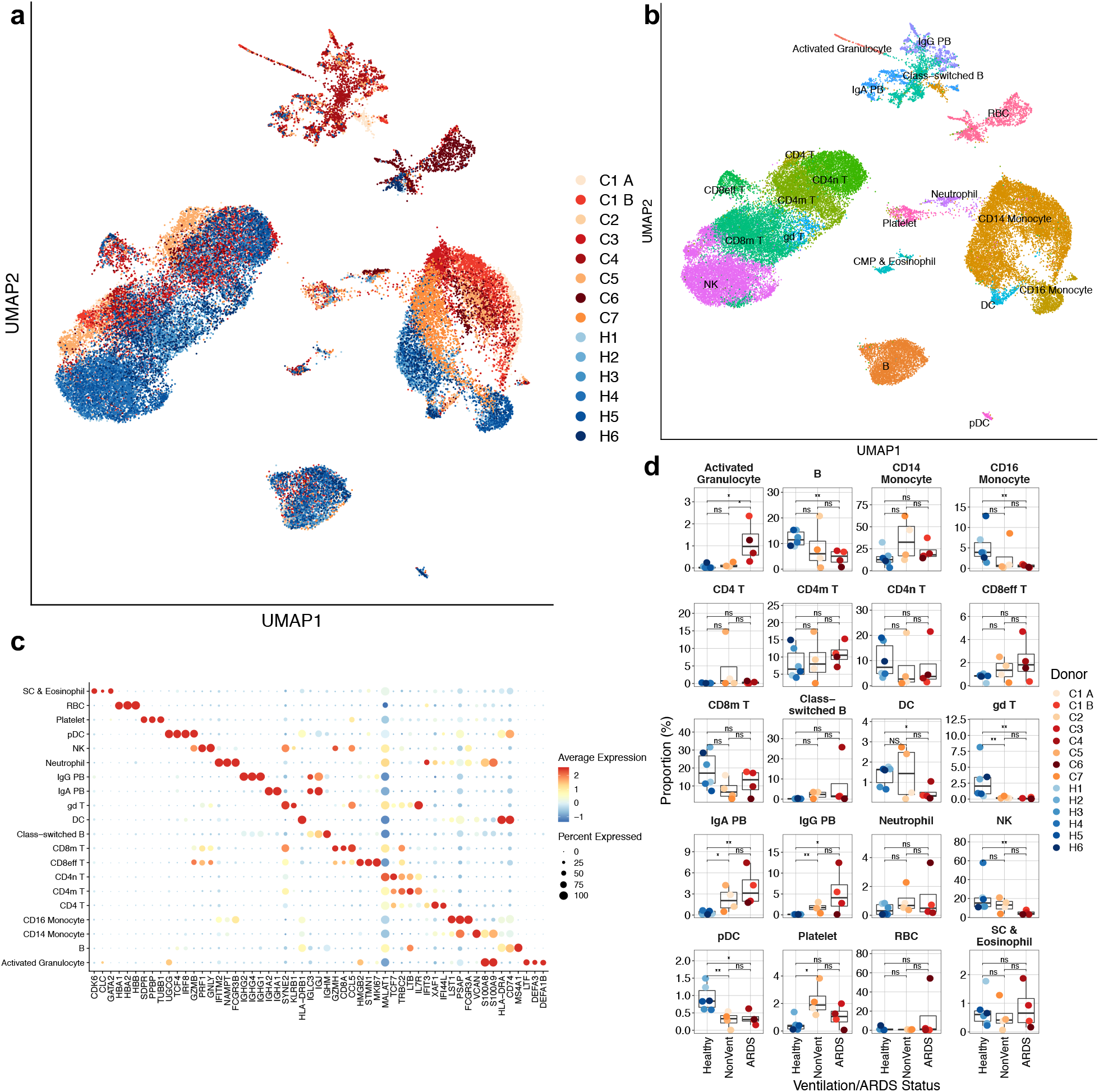
Expansion of plasmablasts and depletion of multiple innate immune cell subsets in the periphery of patients with COVID-19. **a**, UMAP dimensionality reduction embedding of PBMCs from all profiled samples (*n* = 44,721 cells) colored by donor of origin. COVID-19 patient IDs (*n* = 7) begin with “C” and are colored in shades of orange (patients who were not ventilated at time of draw) or red (patients with ARDS who were ventilated at time of draw); healthy donors begin with “H” (*n* = 6) and are colored in blues. **b**, UMAP embedding of the entire dataset colored by orthogonally generated clusters labeled by manual cell type annotation confirmed by SingleR^14^. **c**, Dot plot showing average and percent expression of the 3 most defining genes of each cell type. **d**, Proportions of each cell type in each sample colored by donor of origin. The *x*-axis corresponds to the ventilation or ARDS status of each patient. n.s., not significant; *, p<0.05; **, p<0.01 by Wilcoxon rank sum test.

We next quantified the proportions of all cell types in each sample to identify COVID-19-driven changes in PBMC composition. Several innate immune cell subsets were depleted in patients with COVID-19, including γδ T cells, plasmacytoid dendritic cells (pDCs), conventional dendritic cells (DCs), CD16^+^ monocytes, and NK cells (**Fig. 1d**). Interestingly, the latter three cell types were only significantly depleted in samples from patients with more severe disease with ARDS requiring mechanical ventilation (**Fig. 1d**).

We also noted increased plasmablast proportions in COVID-19 patients, which were further elevated in patients with ARDS (**Table 1, Fig. 1d**), suggesting that more severe cases may be associated with a more robust humoral immune response, similar to previous reports^15,16^. Importantly, there was no correlation between proportions of plasmablasts and days post-symptom onset (**Extended Data Fig. 2**). As plasmablasts are rarely detected in the peripheral blood of healthy patients (**Fig. 1d**), the presence of plasmablasts in every COVID-19 sample analyzed suggests evidence of a SARS-CoV-2 humoral immune response. As plasmablasts express high levels of Ig-encoding mRNAs, we examined if we could detect conserved usage of V gene segments in the plasmablasts of COVID-19 patients. Peripheral plasmablasts from COVID-19 patients did not appear to converge on particular Ig V genes (**Extended Data Fig. 3**).

Lastly, a novel cell population which we annotated as “Activated Granulocytes” was significantly increased only in patients with ARDS (**Fig. 1d**). These cells express several genes encoding neutrophil granule proteins (eg. *ELANE, LTF*, and *MMP8*; see **Fig. 4**), but occupy a similar space as class-switched B cells rather than canonical neutrophils in the UMAP embedding (**Fig. 1b**).

### CD14^+^ monocytes display downregulation of MHC class II and IFN-driven phenotypic reconfiguration

We next analyzed monocytes with more granularity, as this cellular compartment appeared to be the most strongly remodeled in COVID-19 patients (**Fig. 1a**). Dimensionality reduction of monocytes alone indicated a strong phenotypic shift in CD14^+^ monocytes and a depletion of CD16^+^ monocytes (**Fig. 2a, b**). This change was least pronounced in donor C7, the youngest profiled patient with the least severe disease (**Table 1**). We first examined the expression of inflammatory cytokines that have been previously reported to be produced by circulating monocytes in COVID-19^6,7^. Interestingly, we did not identify substantial expression of pro-inflammatory cytokines *TNF, IL6, IL1B, CCL3, CCL4*, or *CXCL2* by peripheral monocytes in any of the profiled cells (**Fig. 2c**). This suggests that circulating monocytes may not contribute to the cytokine storm observed in SARS-CoV-2 infection.

**Figure 2.**
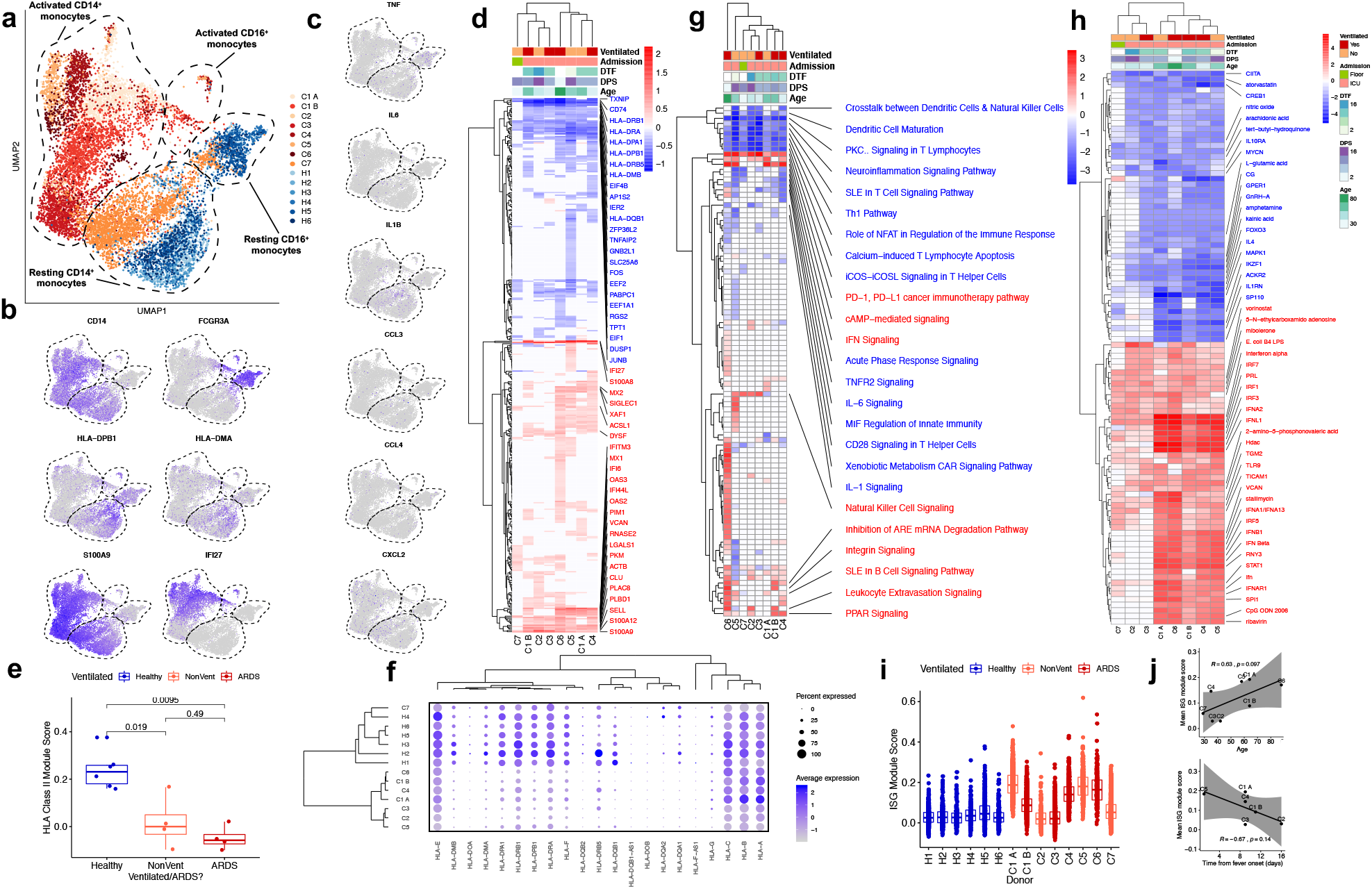
Robust HLA class II downregulation and type I IFN-driven inflammatory signatures in monocytes are characteristics of SARS-CoV-2 infection. **a**, UMAP embedding of all monocytes colored by sample of origin. **b**, UMAP embedding of monocytes colored by *CD14* and *FCGR3A* (CD16, to distinguish between CD14^+^ and CD16^+^ monocytes), *HLA-DPB1* and *HLA-DMA* (illustrating HLA class II downregulation in COVID-19 patients), and *S100A9* and *IFI27* (demonstrating canonical inflammatory signatures in COVID-19 patients). **c**, UMAP embedding of monocytes colored by genes encoding pro-inflammatory cytokines previously reported to be produced by circulating monocytes in severe COVID-19, namely *TNF, IL6, IL1B, CCL3, CCL4*, and *CXCL2*. **d, g, h**, Heatmaps of (**d**) DE genes, (**g**) differentially regulated canonical pathways, and (**h**) differentially regulated predicted upstream regulators between CD14^+^ monocytes of each donor compared to CD14^+^ monocytes of all healthy controls. **d** is colored by average log(fold-change), while **g** and **h** are colored by z-score. All displayed genes, pathways, and regulators are statistically significant at the p<0.05 confidence level. The (**d**) 50 genes, (**g**) 25 pathways, or (**h**) 50 regulators with the highest absolute average log(fold-change) or z-score across all donors all labeled. Genes with a net positive average log(fold-change) or z-score are labeled in red; genes with a net negative average log(fold-change) or z-score are labeled in blue. **e**, Box plot showing the mean HLA Class II module score of CD14^+^ monocytes from each sample, colored by healthy donors (blue), non-ventilated COVID-19 patients (orange), or ventilated COVID-19 patients (red). Shown are exact p values by Wilcoxon rank sum test. **f**, Dot plot depicting percent expression and average expression of all detected *HLA* genes in CD14^+^ monocytes by donor. **i**, Box plot showing the *IFNA* module score of each cell, colored by healthy donors (blue), non-ventilated COVID-19 patients (orange), or ventilated COVID-19 patients (red). **j**, Scatter plots depicting the correlation between the mean ISG module score of each sample and the patient age (top) and time-distance to first measured or reported fever (bottom).

To determine what genes were driving the phenotypic remodelling of monocytes, we identified differentially expressed genes of each COVID-19 sample by comparing cells of each COVID-19 sample to cells of all healthy controls (**Fig. 2d**). We found marked downregulation of 8 genes encoding HLA class II molecules in at least 6 COVID-19 samples relative to healthy controls (**Fig. 2d**). Scoring of all individual cells by their expression of all HLA class II genes revealed that this downregulation was significant in all COVID-19 patients, but potentially more prominent in ventilation-dependent patients (**Fig. 2e**). C7, the patient with the most mild disease course, displayed the least HLA class II downregulation on CD14^+^ monocytes (**Fig. 2f**). HLA class II downregulation is reflected in differentially regulated gene pathways including reduction of “Crosstalk between Dendritic Cells & Natural Killer Cells” and enhancement of “PD-1, PD-L1 cancer immunotherapy pathway” (**Fig. 2g**). Similar HLA class II downregulation was also noted in B cells, where expression of 6 HLA class II genes was significantly lower in at least 6 COVID-19 samples (**Extended Data Fig. 4**). We also noted a trend between HLA class II downregulation and increased age (**Extended Data Fig. 5**). As such downregulation could inhibit the generation of CD4^+^ T cell responses, this could impair adaptive immune responses in older COVID-19 patients. Non-classical HLA class I genes, *HLA-E* and *HLA-F*, were also downregulated to a lesser degree and in fewer samples (**Fig. 2f**), while canonical HLA class I genes *HLA-A, HLA-B*, and *HLA-C* were not consistently up- or down-regulated.

Additionally, 35 type I interferon (IFN)-stimulated genes (ISGs) were upregulated by CD14^+^ monocytes in at least one COVID-19 sample (**Fig. 2d**). Correspondingly, “IFN Signaling” was the second most highly upregulated gene pathway in CD14^+^ monocytes (**Fig. 2g**). Strikingly, this type I IFN signature was not uniform across all COVID-19 samples. Analysis of upstream regulators in CD14^+^ monocytes revealed an absence of predicted IFN and IFN regulatory factor (IRF) activities in donors C2, C3, and C7 relative to the remaining COVID-19 donors (**Figure 2h**). Similar patterns were observed in other cellular compartments (eg. NK cells, see **Fig. 3, Extended Data Figs. 6-10**). To analyze this orthogonally, we scored individual CD14^+^ monocytes in the dataset by their expression of known human type I IFN-stimulated genes and again saw minimal appreciable ISG signatures in donors C2, C3, and C7 (**Fig. 2h**). The differential ISG signature was not explained by ventilation/ARDS (**Fig. 2h, i**), but a higher ISG score trended towards a positive correlation with age and a negative correlation with time-distance to fever onset (**Fig. 2j**). In the patient sampled twice (C1), the ISG module score decreased significantly in the intervening 48 hours between collection of the two samples, during which time the patient decompensated and became ventilator-dependent (**Fig. 2i**). Though causal relationships cannot be established, these data collectively suggest important associations between the peripheral IFN response, patient age, and clinical severity.

**Figure 3.**
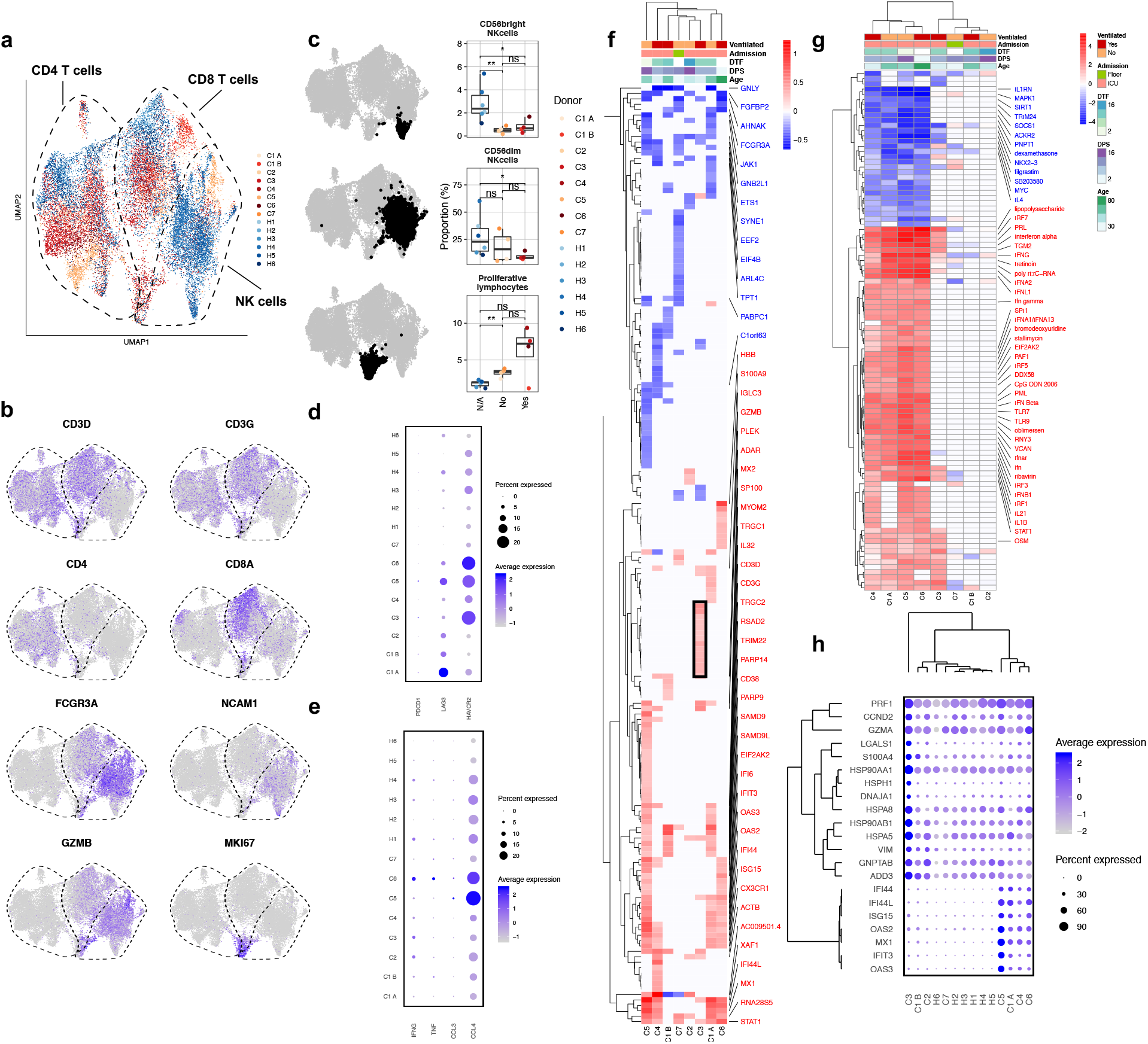
Heterogeneous patterns of NK cell exhaustion and interferon response in COVID-19. **a**, UMAP embedding of CD4^+^ T cells, CD8^+^ T cells, and NK cells colored by sample of origin. **b**, UMAP embedding colored by lineage genes (*CD3D, CD3G, CD4, CD8A, FCGR3A*, and *NCAM1*) and selected functional/phenotypic markers (*GZMB* and *MKI67*). **c**, Box plots depicting proportions of CD56^dim^ NK cells, CD56^bright^ NK cells, and proliferating lymphocytes among total T and NK cells by sample of origin. The cells used to calculate each proportion are highlighted in bold black in the adjacent UMAP embeddings. **, p<0.01; *, p<0.05; n.s., p>0.05 by Wilcoxon rank sum test. **d**, Dot plot showing the percent and average expression of three canonical markers of NK cell exhaustion: *LAG3, PDCD1* (encoding PD-1), and *HAVCR2* (encoding Tim-3). **e**, Dot plot showing the percent and average expression of four canonical NK cell cytokines (*CCL3, CCL4, IFNG*, and *TNF*) by NK cells. **f, g**, Heatmaps of (**f**) DE genes and (**g**) differentially regulated predicted upstream regulators between NK cells of each COVID-19 sample compared to NK cells of all healthy controls. As in **Fig. 2, f** is colored by average log(fold-change), while **g** is colored by z-score. All displayed genes and regulators are statistically significant at the p<0.05 confidence level. The 50 genes or regulators with the highest absolute average log(fold-change) or z-score across all donors all labeled. Genes with a net positive average log(fold-change) or z-score are labeled in red; genes with a net negative average log(fold-change) or z-score are labeled in blue. **f**, Genes that are selectively upregulated by the NK cells of donor C3 are boxed in black. **h**, Dot plot depicting the percent and average expression of C3 NK cell-specific genes boxed in (**f**) as well as selected ISGs upregulated by multiple COVID-19 patients.

### Heterogeneous reconfiguration of peripheral NK cell phenotype in COVID-19

We next analyzed the transcriptomes of T and NK lymphocytes in COVID-19 patient samples as there appeared to be substantial phenotypic shifts in these cellular compartments compared to healthy controls (**Fig. 1a, b**). Indeed, a UMAP embedding of T and NK cells identified substantial differences in the cellular phenotypes of CD4^+^ T, CD8^+^ T, and NK cells (**Fig. 3a, b**). We found that CD56^dim^ NK cells, which are generally thought to contribute to antiviral host defense through cell-mediated cytotoxicity^17^, were depleted primarily in ventilator-dependent patients, whereas CD56^bright^ NK cells, which are considered robust producers of IFN-γ and TNF-α^18^, were significantly depleted in all COVID-19 samples (**Fig. 3c**). Additionally, we identified a cluster of highly proliferative cells, composed of both T and NK cells, that appeared to be increased in most COVID-19 patients (**Fig. 3c**). As SARS-CoV-2 infection has been associated with cytotoxic lymphocyte exhaustion^9^, we next profiled expression of canonical exhaustion markers on CD8^+^ T and NK cells. Surprisingly, there was no clear change in CD4^+^ or CD8^+^ T cell exhaustion in COVID-19 patients (**Extended Data Fig. 11**), but NK cells from COVID-19 patients expressed higher levels of the exhaustion markers *LAG3* and *HAVCR2* compared to healthy controls (**Fig. 3d**).

Similar to our observations in peripheral monocytes, we did not detect substantial expression of pro-inflammatory cytokines by T or NK cells (**Fig. 3e, Extended Data Fig. 12**). Only two patients (C5 and C6) demonstrated substantial upregulation of *CCL4* by peripheral NK cells; all other COVID-19 patients did not express higher levels of *CCL3, CCL4, IFNG*, or *TNF* in CD8^+^ T or NK cells relative to healthy controls (**Fig. 3e, Extended Data Fig. 12**). Additionally, although one group reported a population of GM-CSF producing pathogenic peripheral TH1 cells in COVID-19 by flow cytometry^6^, no reads in our dataset aligned to *CSF2*, which encodes GM-CSF. The absence of pro-inflammatory cytokine expression by peripheral T and NK cells again indicates that peripheral leukocytes may not contribute to cytokine storm in COVID-19.

To further characterize the phenotypes of T and NK cells in COVID-19, we calculated the DE genes from each COVID-19 patient sample relative to all healthy controls, and used these genes to identify enriched gene pathways and upstream regulators. NK cells displayed a remarkably heterogeneous response between COVID-19 patients (**Fig. 3f**). The most frequently downregulated genes included *FCGR3A, AHNAK*, and *FGFBP2*, which are associated with maturity of peripheral NK cells^19^. The most commonly upregulated genes included ISGs and NK cell activation genes like *PLEK* and *CD38*^*20,21*^. We observed a similar heterogeneity of DE genes in CD4^+^ and CD8^+^ T cells, where the most commonly upregulated genes were ISGs (**Extended Data Figs. 8-9**).

Analysis of predicted upstream regulators indicated a strong IFN-driven response that was starkly absent from half of the profiled COVID-19 samples in both NK cells, CD4^+^, and CD8^+^ T cells (**Fig. 3g, Extended Data Figs. 8-10**). Although the most extensive NK cell phenotypic shifts occurred in donors with strong IFN signatures (**Fig. 3f, g**), we identified a group of genes upregulated in patient C3, who had a minimal IFN signature (boxed in **Fig. 3f, Extended Data Figs. 6-10**), that were not upregulated in any other COVID-19 patient. These genes included NK cell cytotoxicity mediators *PRF1* and *GZMB*, as well as 6 heat shock proteins (**Fig. 3h**). A similar group of genes, consisting predominantly of heat shock proteins, was also identified in the CD8^+^ T cells of patient C3 (**Extended Data Fig. 9**). These results collectively indicate that the phenotypes of peripheral T and NK lymphocytes are heterogeneously reconfigured in COVID-19.

### Differentiation of a novel granulocytic cell type from B cells is a characteristic of severe COVID-19 in patients with ARDS

We next analyzed the phenotypes of plasmablasts, class-switched B cells, and activated granulocytes, which appeared to be phenotypically related as they occupied a similar space in a dimensionality reduction embedding of all cells in the dataset (**Fig. 1b**). Indeed, when embedding only these cell types from all donors, activated granulocytes appeared to project linearly from class-switched B cells, suggestive of a continuum of cellular phenotype between the two cell types (**Fig. 4a**). Cellular complexity (the number of genes sequenced per cell divided by the unique molecular identifiers (UMIs) per cell) was not higher in activated granulocytes, making it unlikely that these cells were multiplets (**Extended Data Fig. 13**). To analyze if there was any transition between the two cell types, we performed a cellular trajectory analysis by RNA velocity. RNA velocity analyzes the abundances of spliced and unspliced transcripts to calculate the first derivative of the gene expression state and thus predicts the future state of a cell^22,23^.

**Figure 4.**
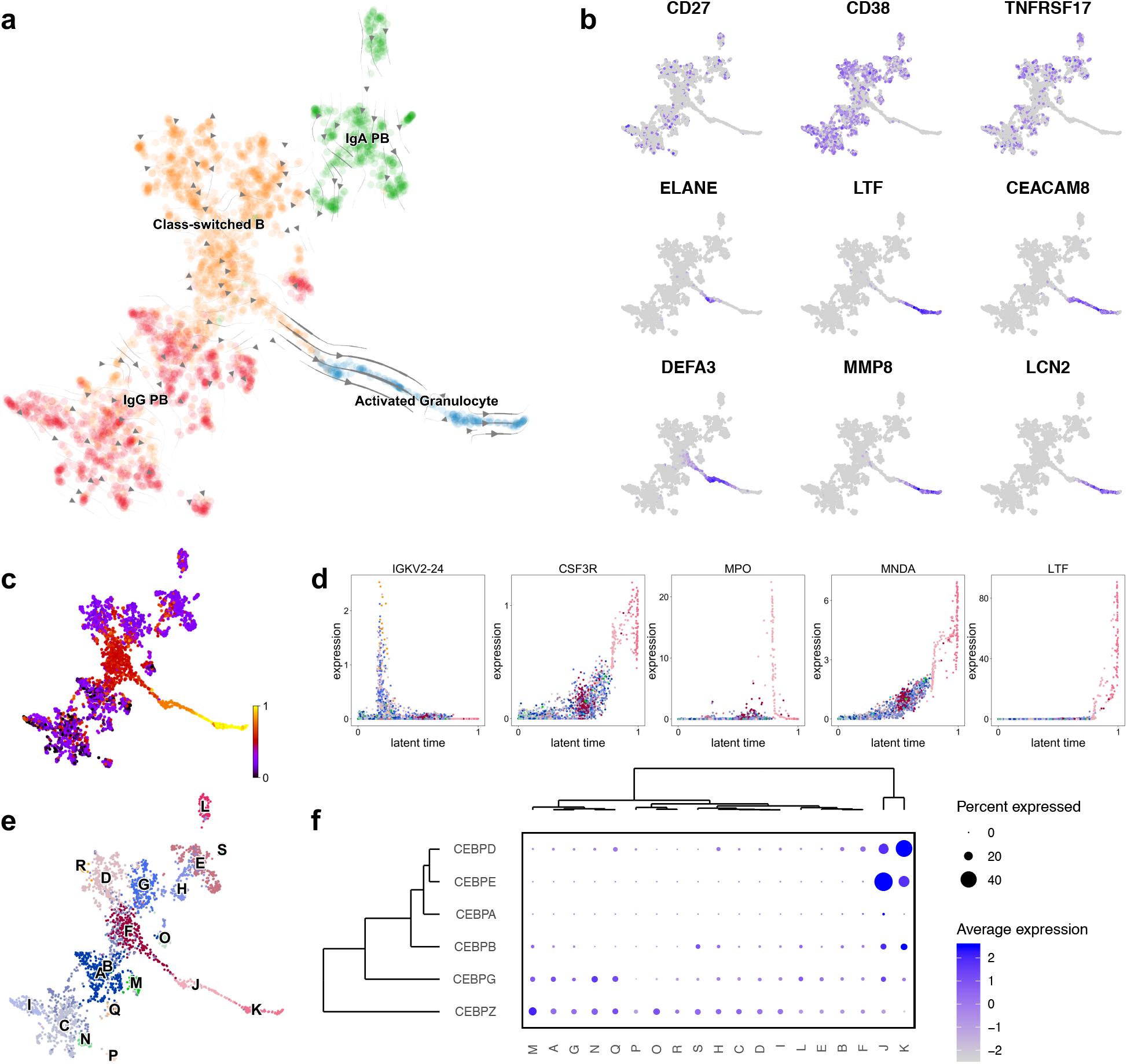
Activated granulocytes are characteristic of severe COVID-19 patients and differentiate from class-switched B cells. **a**, UMAP embedding of plasmablasts, class-switched B cells, and activated granulocytes, colored by annotated cell type and overlaid with RNA velocity stream. **b**, UMAP embedding colored by canonical plasmablast markers (*CD27, CD38*, and *TNFRSF17*) and markers of activated granulocytes (*ELANE, LTF, CEACAM8, DEFA3, MMP8*, and *LCN2*). **c**, UMAP embedding colored by inferred latent time. **d**, Scatter plots showing expression of a selection of cluster-defining genes across inferred latent time. **e**, UMAP embedding colored by orthogonally-generated clusters. **f**, Dot plot depicting expression of *CEBP* family members in each identified cluster.

Surprisingly, this analysis demonstrated that the linear continuum of cellular phenotype represented a differentiation bridge from class-switched B cells to activated granulocytes (**Fig. 4a**). The cells along this differentiation bridge had lost expression of canonical plasmablast markers *CD27, CD38*, and *TNFRSF17* (encoding BCMA) and instead acquired expression of genes encoding neutrophil granule proteins and other granulocytic markers including *ELANE, LTF*, and *DEFA3* (**Fig. 4b**). Recovery of inferred latent time of these cells, which is based solely on a cell’s transcriptional dynamics, also suggested a continuum from class-switched B cells to activated granulocytes (**Fig. 4c**). While cells at the beginning of this continuum are defined by expression of Ig genes, granulocytic markers like *CSF3R* and *MNDA* (encoding myeloid nuclear differentiation antigen) are upregulated as latent time progresses (**Fig. 4d**).

While a lymphocyte-to-granulocyte differentiation process challenges immunological dogma, it is not without precedent. Similar transitions have been described from B cells to macrophages or granulocytes, and the C/EBP transcription factor family has been implicated in controlling this transdifferentiation^24,25^. Interestingly, two C/EBP family members, *CEBPD* and *CEBPE*, are selectively expressed by the two clusters of cells along the differentiation bridge (**Fig. 4e, f**). To identify potential factors that might be driving this transition, we applied NicheNet, a technique that uses gene expression data to discover the putative ligand-target links mediating downstream transcriptional changes^26^. This analysis identified two cytokines, *EGF* and *IL24*, which were both predicted to upregulate *CEBPD* through interaction with *TFRC, IGF2R*, and *M6PR* (for *EGF*), and *NOTCH2* (for *IL24*) (**Extended Data Fig. 14**). There was limited expression of both *EGF* and *IL24* in this dataset, suggesting that if these cytokines were driving the differentiation of activated granulocytes, they may be produced by cells not present in peripheral blood. Collectively, these data suggest a novel transdifferentiation process characteristic of ARDS in severe COVID-19 infection.

## DISCUSSION

We used single cell transcriptomics to characterize the peripheral immune response in severe COVID-19. Overall, we observed marked changes in immune cell composition and phenotype in SARS-CoV-2 infection compared to healthy controls, including depletion of innate immune cells such as NK cells and pDCs, and the induction of an ISG signature across multiple immune cell types. Furthermore, we identified immunological features of severe COVID-19 in patients that required ventilation, including downregulation of HLA class II on monocytes and B cells, and the presence of a population of novel activated granulocytes in peripheral blood. While we found an expansion of plasmablasts in peripheral blood in all COVID-19 patients, we did not detect any conserved Ig V gene usage. B cell repertoire sequencing will be necessary to further analyze the features of SARS-CoV-2 humoral responses.

Downregulation of HLA class II has been observed, both at the transcriptional and the protein level, in patients with severe COVID-19 across multiple studies^27,28^, as well as in SARS-CoV infection^29^. Expression of HLA class II molecules on circulating monocytes mediates antigen presentation to CD4^+^ T cells, and downregulation is associated with decreased responsiveness to stimuli^30^. Downregulation of HLA-DR on circulating monocytes is commonly observed in conditions involving systemic inflammation, including sepsis^31^, and has been correlated to suppressed immune responses, lymphopenia and increased incidence of secondary infections^32,33^. As such, the marked downregulation of HLA class II molecules in COVID-19, particularly in severe disease with ARDS, may reflect immune dysregulation that inhibits the generation of protective immunity. Failure to restore HLA class II expression is associated with poorer outcomes in sepsis, and indeed in our longitudinal patient (C1), a decrease in HLA-DP and HLA-DR expression occurred during worsening clinical course (**Fig. 2d**).

We observed a heterogeneous induction of ISGs in monocytes, T cells and NK cells across all patients, in both mild and severe COVID-19. While interferons are potent mediators of antiviral responses, and have been proposed as a treatment for SARS-CoV-2 infection^34^, they have also been associated with immunopathology in acute viral infections^35^. In SARS-CoV infection, expression of IFNs and ISGs are upregulated at early stages of infection and exhibit a decreased expression over time that is associated with recovery, as persistent ISG induction is associated with poor outcomes^29^. In line with these findings, we also observed an inverse correlation between ISG expression in monocytes with time-distance to fever onset (**Fig. 2j**). A unique characteristic of these SARS-CoV-2-infected patients is that only a subset of donors possessed a robust NK cell ISG signature, and NK cell ISG induction was not associated with ventilation/ARDS status (**Fig. 3f, g**). IFNs promote the activation of antiviral responses in NK cells^36^, and ISGs are upregulated in NK cells in multiple settings of acute viral infection^37^. What differentiates the activation of IFN-mediated pathways in NK cells during SARS-CoV-2 infection, and how this relates to clinical outcomes and the generation of antiviral immunity, remains to be elucidated.

One patient (C1) was sampled longitudinally, once before (C1 A) and after (C1 B) the development of overt ARDS. We observed distinct patterns of immune activation between the two timepoints, which were concordant with the findings that distinguished the patients with ARDS in the remaining subjects. This provides insight into the highly dynamic peripheral immune response during progression to ARDS in SARS-CoV-2 infection. While patient C1 initially exhibited a strong immune activation signature, including expression of ISG modules and cytotoxic T and NK cell exhaustion, this was significantly dampened after the onset of ARDS. This could reflect the role of IFN-mediated responses in ARDS, where excessive immune activation and IFN signaling may lead to lung inflammation and immunopathology^38^.

Our analyses also revealed a population of activated granulocytes present in peripheral blood only in severe disease with ARDS (**Fig. 1d**). These cells are not likely to represent granulocytes that have phagocytosed B cells, a feature of hemophagocytic lymphohistiocytosis (HLH) which can be triggered by severe acute infections, because these patients did not have other characteristics of HLH such as markedly elevated ferritin. Additionally, cells do not appear to be doublets, because the complexity (the number of genes per UMI detected) of these cells was comparable to that of other cell types. Instead, we show that these cells likely transdifferentiate from class-switched B cells (**Fig. 4**) and express transcription factors of the C/EBP family including *CEBPD* and *CEBPE*, both known inducers of myeloid and granulocyte cell fates^39,40^. The reprogramming of murine B cells via the exogenous expression of C/EBPs has demonstrated the capability of overriding B cell lineage commitment to allow transdifferentiation into granulocytes^25^. While we did not identify expression of upstream drivers of C/EBP induction in peripheral blood (**Extended Data Fig. 13**), this transdifferentiation process may be induced by signals from the lungs in SARS-CoV-2 infected patients. Indeed, murine studies of influenza infection have found a strong transcriptional upregulation of IL-24 in lungs post-infection^41^. *In vitro* infection of human lung epithelial cells with the same virus also induces IL-24 production^42^, suggesting that infection-driven production of IL-24 can act as a potential signal to induce this novel transdifferentiation.

Increased neutrophil counts have been reported in patients with severe COVID-19^4,16,43^. We analyzed PBMCs, which generally exclude neutrophils during purification, but of those that remained, we did not observe a significant increase in the proportion of canonical neutrophils within our samples (**Fig. 1d**). This suggests that the increase in neutrophils observed clinically may be due to the increase of this activated granulocyte subset. The role of granulocytes in COVID-19 pathogenesis will be an important area of future study.

In summary, we have presented a single-cell atlas of the peripheral immune response to COVID-19. These data highlight immunological features associated with severe disease, and can inform therapeutic strategies via an improved understanding of mechanisms of immune dysregulation.

## MATERIALS & METHODS

### Subjects and specimen collection

We collected blood from 7 patients enrolled in the Stanford University ICU Biobank study from March-April 2020 after written informed consent from patients or their surrogates (Stanford IRB approval #28205). Eligibility criteria included age > = 18 years and admission to Stanford Hospital with a positive SARS-CoV-2 nasopharyngeal swab by RT-PCR. Patients admitted to the wards or ICU were included, and the majority were co-enrolled in ongoing COVID-19 treatment trials at Stanford. Screening of new admissions via electronic medical records review of all subjects was performed by the study coordinator (JR), research fellow (AR), COVID-19 clinical consultants (PG and AS), and the study PI (AJR), and was done every day with a goal enrollment in <48 h of admission to the hospital. Patients were phenotyped for ARDS using Berlin criteria (acute onset of hypoxemic respiratory failure with a PaO_2_/FIO_2_ ratio < 300 on at least 5cm of PEEP, bilateral infiltrates on Chest X-Ray)^44^. In order to protect the identity of the COVID-19 subjects, ages are reported as ranges. For controls, blood was collected from 6 healthy adult donors as part of the Profiling Healthy Immunity study after written informed consent (Stanford IRB approval #26571). All donors were consented for genetic research.

For patients, blood draws occurred in concert with usual care to avoid unnecessary personal protective equipment usage. Blood was collected into heparin tubes (Becton, Dickinson, and Co.), and PBMCs were isolated by density gradient centrifugation using Ficoll-Paque PLUS (GE Healthcare) and washed with Ca/Mg-free PBS. Processing of blood began within four hours of collection for all samples, and within one hour for most.

### Single-cell RNA sequencing by Seq-Well

The Seq-Well platform for scRNA-seq was utilized as described previously^13,45^. Immediately after Ficoll separation, 50,000 PBMCs were resuspended in RPMI + 10% FCS at a concentration of 75,000 cells/mL. 200 μL of this cell suspension (15,000 cells) was then loaded onto Seq-Well arrays pre-loaded with mRNA capture beads (ChemGenes). Following four washes with DPBS to remove serum, the arrays were sealed with a polycarbonate membrane (pore size of 0.01 µm) for 30 minutes at 37°C and then frozen at -80°C for no less than 24 hours and no more than 14 days to allow batching of samples processed at irregular hours. Next, arrays were thawed, cells lysed, transcripts hybridized to the mRNA capture beads, and beads recovered from the arrays and pooled for downstream processing. Immediately after bead recovery, mRNA transcripts were reverse transcribed using Maxima H-RT (Thermo Fisher EPO0753) in a template-switching-based RACE reaction, excess unhybridized bead-conjugated oligonucleotides removed with Exonuclease I (NEB M0293L), and second-strand synthesis performed with Klenow fragment (NEB M0212L) to enhance transcript recovery in the event of failed template switching^45^. Whole transcriptome amplification (WTA) was performed with KAPA HiFi PCR Mastermix (Kapa Biosystems KK2602) using approximately 6,000 beads per 50 μL reaction volume. Resulting libraries were then pooled in sets of 6 (approximately 36,000 beads per pool) and products purified by Agencourt AMPure XP beads (Beckman Coulter, A63881) with a 0.6x volume wash followed by a 0.8x volume wash. Quality and concentration of WTA products was determined using an Agilent Fragment Analyzer (Stanford Functional Genomics Facility), with a mean product size of >800bp and a non-existent primer peak indicating successful preparation. Library preparation was performed with a Nextera XT DNA library preparation kit (Illumina FC-131-1096) with 1 ng of pooled library using dual-index primers. Tagmented and amplified libraries were again purified by Agencourt AMPure XP beads with a 0.6x volume wash followed by a 1.0x volume wash, and quality and concentration determined by Fragment Analysis. Libraries between 400-1000bp with no primer peaks were considered successful and pooled for sequencing. Sequencing was performed on a NovaSeq S2 instrument (Illumina; Chan Zuckerberg Biohub). The read structure was paired-end with read 1 beginning from a custom read 1 primer^13^ containing a 12bp cell barcode and an 8 bp unique molecular identifier (UMI), and with read 2 containing 50bp of mRNA sequence.

### Alignment and quality control of sequencing data

Sequencing reads were aligned and count matrices assembled using STAR^46^ and dropEst^47^, respectively. Briefly, the mRNA reads in read 2 demultiplexed FASTQ files were tagged with the cell barcode and UMI for the corresponding read in the read 1 FASTQ file using the dropTag function of dropEst. Next, reads were aligned with STAR using the GRCh37 (hg19) human reference genome that included the complete genome sequences for all SARS-CoV-2 strains sequenced from California before March 24, 2020 (10 SARS-CoV-2 sequences). No SARS-CoV-2 reads were aligned from these samples using this strategy, even when the outFilterMultimapNmax behavioral option of STAR was increased from 10 (default) to 20 to accommodate for potential multiple-mapping SARS-CoV-2 reads. Count matrices were built from resulting BAM files using dropEst^47^. Count matrices for intron-aligned reads were also generated in order to computationally analyze cellular trajectory. Cells that had fewer than 1,000 UMIs or greater than 15,000 UMIs, as well as cells that contained greater than 20% of reads from mitochondrial genes or rRNA genes (*RNA18S5* or *RNA28S5*), were considered low quality and removed from further analysis. To remove putative multiplets (where more than one cell may have loaded into a given well on an array), cells that expressed more than 75 genes per 100 UMIs were also filtered out. Genes that were expressed in fewer than 10 cells were removed from the final count matrix.

### scRNA-seq computational pipelines and analyses

The R package Seurat was used for data scaling, transformation, clustering, dimensionality reduction, differential expression analysis, and most visualization^48^. Data were scaled and transformed and variable genes identified using the SCTransform() function, and linear regression performed to remove unwanted variation due to cellular complexity (# of genes per cell, # of UMIs per cell) or cell quality (% mitochondrial reads, % rRNA reads). PCA was performed using variable genes, and the first 50 principal components (PCs) used to perform UMAP to embed the dataset into two dimensions. Next, the first 50 PCs were used to construct a shared nearest neighbor graph (SNN; FindNeighbors()) and this SNN used to cluster the dataset (FindClusters()). Despite upstream filtering for high quality cells and regression on gene’s reflective of cell quality, 2 clusters were identified where 65% or 100% of the positively enriched genes were of mitochrondrial or ribosomal origin, and these clusters were removed from further analysis^49,50^. Cellular identity was determined by finding differentially expressed genes for each cluster using Seurat’s implementation of the Wilcoxon rank-sum test (FindMarkers()) and comparing those markers to known cell type-specific genes from previous datasets^51–56^. Cluster annotation was confirmed using the R package SingleR^14^, which compares the transcriptome of each single cell to reference datasets in order to determine cellular identity. The majority of cells in cluster 24 were labeled as common myeloid progenitors by SingleR, but this cluster also contained cells annotated as 7 different lineages of hematopoietic stem cells and progenitors. Closer examination revealed that this cluster consisted of two groups of cells, one which expressed *CLC* and the other which expressed *CD34*, and we therefore labeled them as stem cells (SC) & eosinophils for downstream analysis. 98% of cells in cluster 27 were annotated by SingleR as Myelocytes (46%), Pro-Myelocytes (22%), CD34^-^ pre-B cells (14%), or HSC G-CSF (17%). However, these cells expressed several genes encoding for neutrophil granule proteins (eg. *ELANE, MPO, LTF, CTSG, LCN2*, and *MMP8*) yet were distinct from cluster 25 (labeled manually and by SingleR as neutrophils) and did not express canonical neutrophil markers like *FCGR3B* and *CXCR2*. Thus, we annotated these cells as “Activated Granulocytes”.

Gene pathway and upstream regulator analysis was performed with Ingenuity Pathway Analysis (IPA; Qiagen). The parent Seurat object was divided into individual objects consisting of cells from a particular cellular compartment (eg. CD4^+^ T cells, NK cells, CD16^+^ Monocytes, etc.). Next, differentially expressed genes between the cells from each COVID-19 patient sample and the cells from all healthy controls were calculated by FindMarkers(). The average log(fold change) of each DE gene calculated by FindMarkers() was supplied to IPA. Modeling of intracellular communication was performed by NicheNet^26^, using activated granulocytes and the DE genes of activated granulocytes as the receiver cell population and the geneset of interest, respectively. Analysis of cellular trajectory by RNA velocity was performed using the package scVelo using dynamical modeling^23^. Dot plots with hierarchical clustering were generated using FlexDotPlot^57^.

## Data Availability

Raw sequencing data will be deposited on GEO. Processed count matrices will be hosted on and available for download from the publicly accessible cellxgene platform by the Chan Zuckerberg Biohub Initiative.

## ACKNOWLEDGEMENTS

We are grateful to all participants in this study. We thank Sopheak Sim and the Stanford Functional Genomics Facility for the use of the Fragment Analyzer, as well as Angela Detweiler and Michelle Tan at the Chan Zuckerberg Biohub for assistance with sequencing.

The Stanford ICU Biobank and A.J.R. are funded by NIH/NHLBI K23 HL125663. A.J.W. is supported by the Stanford Medical Scientist Training Program (T32 GM007365-44) and the Stanford Bio-X Interdisciplinary Graduate Fellowship; A.R. is supported by the Applied Genomics in Infectious Diseases training grant T32 AI007502-23; N.Q.Z. is supported by a National Science Scholarship from A*STAR Singapore; J.L.M. is supported by National Science Foundation Graduate Research Fellowship DGE-1656518; J.L.M. and G.I. are supported by NIH training grant T32 AI007290-35. C.A.B. is supported by NIH/NIDA DP1 DA04608902, a 2019 Sentinel Pilot Project from the Bill & Melinda Gates Foundation, and Burroughs Wellcome Fund Investigators in the Pathogenesis of Infectious Diseases #1016687. C.A.B. is the Tashia and John Morgridge Faculty Scholar in Pediatric Translational Medicine from the Stanford Maternal Child Health Research Institute and an Investigator of the Chan Zuckerberg Biohub.

## AUTHOR CONTRIBUTIONS

C.A.B. and A.J.R. conceived the study. A.S., P.G., A.J.R., J.R. and A.R. identified eligible patients. A.J.R., A.R. and J.R. enrolled and consented patients. A.J.W., N.Q.Z., A.R., G.J.M., J.L.M., G.T.I., T.R., R.V., T.H. and L.J.S. processed patient samples. A.J.W and N.Q.Z. performed scRNA-seq and computational analysis. A.J.W., A.R., N.Q.Z. and C.A.B. interpreted data and wrote the manuscript with input from all authors.

## REFERENCES

1. Wu, Z. & McGoogan, J. M. Characteristics of and Important Lessons From the Coronavirus Disease 2019 (COVID-19) Outbreak in China: Summary of a Report of 72 314 Cases From the Chinese Center for Disease Control and Prevention. JAMA (2020) doi:10.1001/jama.2020.2648.

2. CDC COVID-19 Response Team. Severe Outcomes Among Patients with Coronavirus Disease 2019 (COVID-19) - United States, February 12-March 16, 2020. MMWR Morb. Mortal. Wkly. Rep. 69, 343–346 (2020).

3. Guan, W.-J. et al. Clinical Characteristics of Coronavirus Disease 2019 in China. N. Engl. J. Med. (2020) doi:10.1056/NEJMoa2002032.

4. Huang, C. et al. Clinical features of patients infected with 2019 novel coronavirus in Wuhan, China. Lancet 395, 497–506 (2020).

5. Chen, G. et al. Clinical and immunologic features in severe and moderate Coronavirus Disease 2019. J. Clin. Invest. (2020) doi:10.1172/JCI137244.

6. Zhou, Y. et al. Pathogenic T cells and inflammatory monocytes incite inflammatory storm in severe COVID-19 patients. Natl Sci Rev (2020) doi:10.1093/nsr/nwaa041.

7. Guo, C. et al. Tocilizumab treatment in severe COVID-19 patients attenuates the inflammatory storm incited by monocyte centric immune interactions revealed by single-cell analysis. bioRxiv 2020.04.08.029769 (2020) doi:10.1101/2020.04.08.029769.

8. Zheng, H.-Y. et al. Elevated exhaustion levels and reduced functional diversity of T cells in peripheral blood may predict severe progression in COVID-19 patients. Cell. Mol. Immunol. (2020) doi:10.1038/s41423-020-0401-3.

9. Diao, B. et al. Reduction and Functional Exhaustion of T Cells in Patients with Coronavirus Disease 2019 (COVID-19). Infectious Diseases (except HIV/AIDS) (2020) doi:10.1101/2020.02.18.20024364.

10. Xu, Z. et al. Pathological findings of COVID-19 associated with acute respiratory distress syndrome. Lancet Respir Med 8, 420–422 (2020).

11. Peiris, J. S. M. et al. Clinical progression and viral load in a community outbreak of coronavirus-associated SARS pneumonia: a prospective study. Lancet 361, 1767–1772 (2003).

12. Nicholls, J. M. et al. Lung pathology of fatal severe acute respiratory syndrome. Lancet 361, 1773–1778 (2003).

13. Gierahn, T. M. et al. Seq-Well: portable, low-cost RNA sequencing of single cells at high throughput. Nat. Methods 14, 395–398 (2017).

14. Aran, D. et al. Reference-based analysis of lung single-cell sequencing reveals a transitional profibrotic macrophage. Nat. Immunol. 20, 163–172 (2019).

15. Zhao, J. et al. Antibody responses to SARS-CoV-2 in patients of novel coronavirus disease 2019. Clin. Infect. Dis. (2020) doi:10.1093/cid/ciaa344.

16. Zhang, B. et al. Immune phenotyping based on neutrophil-to-lymphocyte ratio and IgG predicts disease severity and outcome for patients with COVID-19. Infectious Diseases (except HIV/AIDS) (2020) doi:10.1101/2020.03.12.20035048.

17. Caligiuri, M. A. Human natural killer cells. Blood 112, 461–469 (2008).

18. Poli, A. et al. CD56bright natural killer (NK) cells: an important NK cell subset. Immunology 126, 458–465 (2009).

19. Hydes, T. et al. IL-12 and IL-15 induce the expression of CXCR6 and CD49a on peripheral natural killer cells. Immun Inflamm Dis 6, 34–46 (2018).

20. Smith, S. L. et al. Diversity of peripheral blood human NK cells identified by single-cell RNA sequencing. Blood Adv 4, 1388–1406 (2020).

21. Gars, M. L. et al. Pregnancy-Induced Alterations in NK Cell Phenotype and Function. Frontiers in Immunology vol. 10 (2019).

22. La Manno, G. et al. RNA velocity of single cells. Nature 560, 494–498 (2018).

23. Bergen, V., Lange, M., Peidli, S., Alexander Wolf, F. & Theis, F. J. Generalizing RNA velocity to transient cell states through dynamical modeling. bioRxiv 820936 (2019) doi:10.1101/820936.

24. Xie, H., Ye, M., Feng, R. & Graf, T. Stepwise Reprogramming of B Cells into Macrophages. Cell vol. 117 663–676 (2004).

25. Cirovic, B. et al. C/EBP-Induced Transdifferentiation Reveals Granulocyte-Macrophage Precursor-like Plasticity of B Cells. Stem Cell Reports 8, 346–359 (2017).

26. Browaeys, R., Saelens, W. & Saeys, Y. NicheNet: modeling intercellular communication by linking ligands to target genes. Nat. Methods 17, 159–162 (2020).

27. Ong, E. Z., et al. A dynamic immune response shapes COVID-19 progression. Cell Host Microbe doi:10.1016/j.chom.2020.03.021.

28. Giamarellos-Bourboulis, E. J., et al. Complex Immune Dysregulation in COVID-19 Patients with Severe Respiratory Failure. Cell Host Microbe doi:10.1016/j.chom.2020.04.009.

29. Cameron, M. J. et al. Interferon-mediated immunopathological events are associated with atypical innate and adaptive immune responses in patients with severe acute respiratory syndrome. J. Virol. 81, 8692–8706 (2007).

30. Veglia, F., Perego, M. & Gabrilovich, D. Myeloid-derived suppressor cells coming of age. Nat. Immunol. 19, 108–119 (2018).

31. Le Tulzo, Y. et al. Monocyte human leukocyte antigen-DR transcriptional downregulation by cortisol during septic shock. Am. J. Respir. Crit. Care Med. 169, 1144–1151 (2004).

32. Döcke, W. D. et al. Monocyte deactivation in septic patients: restoration by IFN-gamma treatment. Nat. Med. 3, 678–681 (1997).

33. Landelle, C. et al. Low monocyte human leukocyte antigen-DR is independently associated with nosocomial infections after septic shock. Intensive Care Med. 36, 1859–1866 (2010).

34. Sallard, E., Lescure, F.-X., Yazdanpanah, Y., Mentre, F. & Peiffer-Smadja, N. Type 1 interferons as a potential treatment against COVID-19. Antiviral Res. 178, 104791 (2020).

35. Teijaro, J. R. Type I interferons in viral control and immune regulation. Curr. Opin. Virol. 16, 31–40 (2016).

36. Paolini, R., Bernardini, G., Molfetta, R. & Santoni, A. NK cells and interferons. Cytokine Growth Factor Rev. 26, 113–120 (2015).

37. Boeijen, L. L., Hou, J., de Groen, R. A., Verbon, A. & Boonstra, A. Persistent Replication of HIV, Hepatitis C Virus (HCV), and HBV Results in Distinct Gene Expression Profiles by Human NK Cells. Journal of Virology vol. 93 (2018).

38. Makris, S., Paulsen, M. & Johansson, C. Type I Interferons as Regulators of Lung Inflammation. Front. Immunol. 8, 259 (2017).

39. Lekstrom-Himes, J. A. The role of C/EBP(epsilon) in the terminal stages of granulocyte differentiation. Stem Cells 19, 125–133 (2001).

40. Balamurugan, K. & Sterneck, E. The many faces of C/EBPδ and their relevance for inflammation and cancer. Int. J. Biol. Sci. 9, 917–933 (2013).

41. Park, S.-J. et al. Dynamic changes in host gene expression associated with H5N8 avian influenza virus infection in mice. Sci. Rep. 5, 16512 (2015).

42. Seong, R.-K., Choi, Y.-K. & Shin, O. S. MDA7/IL-24 is an anti-viral factor that inhibits influenza virus replication. J. Microbiol. 54, 695–700 (2016).

43. Qin, C. et al. Dysregulation of immune response in patients with COVID-19 in Wuhan, China. Clin. Infect. Dis. (2020) doi:10.1093/cid/ciaa248.

44. ARDS Definition Task Force et al. Acute respiratory distress syndrome: the Berlin Definition. JAMA 307, 2526–2533 (2012).

45. Hughes, T. K. et al. Highly Efficient, Massively-Parallel Single-Cell RNA-Seq Reveals Cellular States and Molecular Features of Human Skin Pathology. bioRxiv 689273 (2019) doi:10.1101/689273.

46. Dobin, A. et al. STAR: ultrafast universal RNA-seq aligner. Bioinformatics 29, 15–21 (2013).

47. Petukhov, V. et al. dropEst: pipeline for accurate estimation of molecular counts in droplet- based single-cell RNA-seq experiments. Genome Biol. 19, 78 (2018).

48. Butler, A., Hoffman, P., Smibert, P., Papalexi, E. & Satija, R. Integrating single-cell transcriptomic data across different conditions, technologies, and species. Nat. Biotechnol. 36, 411–420 (2018).

49. Carter, R. A. et al. A Single-Cell Transcriptional Atlas of the Developing Murine Cerebellum. Curr. Biol. 28, 2910–2920.e2 (2018).

50. Freytag, S., Tian, L., Lönnstedt, I., Ng, M. & Bahlo, M. Comparison of clustering tools in R for medium-sized 10x Genomics single-cell RNA-sequencing data. F1000Res. 7, 1297 (2018).

51. Gutierrez-Arcelus, M. et al. Lymphocyte innateness defined by transcriptional states reflects a balance between proliferation and effector functions. Nat. Commun. 10, 687 (2019).

52. Villani, A.-C. et al. Single-cell RNA-seq reveals new types of human blood dendritic cells, monocytes, and progenitors. Science 356, (2017).

53. Kang, H. M. et al. Multiplexed droplet single-cell RNA-sequencing using natural genetic variation. Nat. Biotechnol. 36, 89–94 (2018).

54. Nimmerjahn, F. & Ravetch, J. V. Fcgamma receptors as regulators of immune responses. Nat. Rev. Immunol. 8, 34–47 (2008).

55. Nimmerjahn, F. & Ravetch, J. V. Fcgamma receptors: old friends and new family members. Immunity 24, 19–28 (2006).

56. Palmer, C., Diehn, M., Alizadeh, A. A. & Brown, P. O. Cell-type specific gene expression profiles of leukocytes in human peripheral blood. BMC Genomics 7, 115 (2006).

57. Leonard, S., Rolland, A., Tarte, K., Chalmel, F. & Lardenois, A. FlexDotPlot: a universal and modular dot plot visualization tool for complex multifaceted data. bioRxiv 2020.04.03.023655 (2020) doi:10.1101/2020.04.03.023655.

